# Mind the Gap! Developing Patient Responsive Information for Epilepsy

**DOI:** 10.1101/2022.08.23.22279110

**Authors:** Sarah Jones, Heather Angus-Leppan

## Abstract

Technology must adapt to changing expectations about shifting patient needs and accessing health services. Standardised websites provide information often in the “frequently asked questions” format. There is little research about how much these answer the questions of people with epilepsy (PWE).

This study used social media data to analyse the questions PWE have relating to “what would you ask a neurologist/epileptologist” and “do you have any questions relating to your epilepsy”. The study resulted in 2752 questions from PWE in Europe, North America and Australia, presented in the raw data format of natural language. Questions were themed using an unsupervised topic modelling algorithm to process and categorise the data into an aggregated question set.

Many of the questions are not currently answered by Epilepsy charity and medical websites, and many centre on restrictions and fears about lifestyle. This study acts as the first stage toward the supervised topic classification: providing a list of questions to be submitted for answering by healthcare professionals required for a Virtual Assistant.

The ultimate aim of the project is to generate a Virtual Assistant/Chatbot for the use of PWE to provide accurate and interactive responses to their real questions.

## Introduction

Meeting patient needs starts with establishing what they are, and how they change. The generations now moving through the healthcare system require technology to match their needs in terms of interoperability, portability and immediacy (1). Expectations about accessing health services and patient needs are shifting.

Healthcare is increasingly technological; solutions include Electronic Health Records, Clinical Decision Making systems and Remote Patient Monitoring applications (2). As the world moves through and beyond the COVID19 pandemic, the ongoing impact upon healthcare is likely to be significant, particularly for patients with disabilities and/or chronic conditions (3). Technology as a vehicle to meet patients’ needs has great potential, assuming that actual needs are met.

Peer support is an established and effective method for linking people with a health condition, promoting self-management (4), a discussion forum (5) and reducing stigma through the use of the concept of personal/patient empowerment (6). Feeling included into a community, increasing positive self-image and information access through the use of shared, lived experience is key to the interventions covered by the concept of empowerment; although there is certainly merit in the further investigation as to whether this is truly empowerment or a preferable terminology should be sought (7).

Technology, via social media and community platforms, has enabled peer support groups to reach a global audience, decreasing social isolation and increasing the spread of education and information to more diverse and isolated communities. The issues facing People with Epilepsy (PWE), exacerbated by the pandemic (8), stem from the effect of this chronic condition on all areas of life; social, employment, education and relationships. Multiple platforms have been constructed to provide peer support to PWE through community, social and visual media where questions and advice are freely shared.

With this increase in access and amount information comes the associated risk of misinformation and a new problem: how do PWE sift through the amount of information and assess which is correct? Machine learning assists in verifying “fake news” uploaded to platforms (9), however when information gains momentum through word of mouth, peer support groups and social media, a real danger to patient safety can be presented (10).

During a review of common online epilepsy peer support platforms (MyEpilepsyTeam, Brain Ablaze, Facebook groups and #epilepsy threads in Twitter, Instagram and TikTok), it became apparent that people with epilepsy require advice which is not being addressed through media supplied for patient use by healthcare/clinical services. Lived experience accounts detail the difficulties with asking questions at appointments: embarrassment (11), focus on clinical issues (12), not wanting to “waste clinicians time” (13), not having a relationship with the clinician (14), and forgetting to ask/lack of understanding (15). A virtual platform provides a forum to pose questions when they arise, using an anonymous format.

On the surface the peer support method appears to be filling a gap, providing the information patients need. It is important that knowledge comes from appropriate clinical or advisory sources (16). Social media and peer support presents a potential danger to patients if answers are provided through reported experience or hearsay without verification.

Charities and medical organisations provide a range of advice and FAQ sheets. Answers on FAQs are written to accommodate generic needs: whether carer or patient, clinical or “social” questions, in a format which can be easily read. Initially these were on paper so limited to at most to two A4 sides of brief answers.

With the rapid development of mobile devices and the Internet, information dissemination has evolved from printed media and television or radio broadcasting, to mobile devices and applications. To ascertain the current situation relating to advice for PWE a search was undertaken, using Google on Chrome to search the term “Epilepsy Advice”.

### The top results are as shown

This table illustrates that the current method for providing information to PWE is a simple digital conversion, transferring traditional “paper FAQ” into a webpage. Only one website provided the information in video format in addition to text when this study was undertaken.

The results of this search presents questions relating to the ability for patients to find the correct information. Search Engines present information in a ranking basis of relevance; where relevance is subjective, based on algorithms (17), leading to issues of geographical location and poor specificity of answer. There are related issues due to regional application of language /natural language processing, poor queries, or that the information required is so specific that it cannot be found. The information provided is generic, wide in scope, non-specific and not applicable to all age ranges or demographics.

When information is beyond the scope of the FAQ, patients and carers are usually directed to a telephone helpline or email. The use of the telephone helpline, whilst seemingly a suitable mitigation to the issues of text, conflicts with the move of society towards technology; younger members of society preferring non-verbal communication (18), a situation exacerbated by co-morbidities or socio-economic status (19).

The status quo is a necessity of the text web-based format: health information is written in language that could be understood by a typical 12-year-old, although one in every five adults finds this difficult (20). To ensure maximum understanding, information is presented succinctly with a minimum of bulk text.

The FAQ format assumes an understanding has been gained of the information patients require. Where patients are consulted, there will be a risk of limited demography; whether due to time constraints, fear or lack of interest etc (21). There will be an inherent bias of those co-ordinating and writing the information as to how they interpret the needs of patients, balancing these with the information that clinicians deem a priority (22). Such a mismatch in interpretation poses the question as to whether there is a clear understanding of the true concerns of people with epilepsy.

Patient centred care can be summarised as knowing what matters to the patient rather than what is the matter with the patient. (23) It potentially opens clear communication lines, informs for shared decision and increases adherence to medication, patient satisfaction and improves outcomes (24). Due to the increased risk of mental health effects, social exclusion and reduction in self-esteem and confidence in people with epilepsy (25) it is crucial to understand the concerns of patients: finding methods to answer questions with timely and accurate information, reducing the risk of inaccurate information taking precedence thus mitigating the primacy effect (26).

The aim of this study was to undertake the first stages toward using familiar, accessible technology to answer the concerns and queries of PWE and to test a proposed solution

The proposed vehicle is that of a conversational virtual assistant, or chatbot. Users interact with a chat interface, asking questions using natural language. The aim of the virtual assistants is to develop “human like” capabilities of speech and language detection (26) using artificial intelligence. The user remains anonymous to the virtual assistant, with the intention of removing the barriers of embarrassment or other emotions (25).

Once constructed, virtual assistants provide a log of all questions asked/answered through which analysis can determine the current needs of PWE at any given point in time. When a question cannot be answered, it is stored in the log files to be answered by the development team. The hypothesis is that the data should give a true reflection, over time, of the true concerns facing people with epilepsy. By extrapolation, changes in the subject of question should reflect the success of communication/advertising strategies. The nature of a virtual assistant as an online entity facilitates the provision of information to patients and clinicians, in areas where there is little clinical support for epilepsy.

## Method: Review of Existing Datasets

Virtual Assistants require an input of data into a question database; the questions that may be asked (intents), the “search tags” and the answers. The first build stage is to establish a list of questions that may be asked in addition to those provided by current FAQs. Through reviewing public secondary data of questions from online epilepsy support providers, peer support platforms and social media, a list of the most frequently asked questions was established to understand the needs of PWE accessing peer support.

The data reviewed related to answers to the questions “what would you ask a neurologist/epileptologist” and “do you have any questions relating to your epilepsy”. Data relating to other question variants was not considered. The datasets compiled consisted of a question, the country of origin and role (PWE/Clinician/Carer) of the person asking the question (if provided).

There was no data relating to age/gender or other demographics. Due to the anonymity of the data, it was not possible to ascertain whether the questions pertained to individuals asking singular questions or less asking multiple questions. These were not perceived to be limitations as the focus of the review was to ascertain a question database/dataset for use in the virtual assistant but may be a useful consideration for further research into the needs and concerns of specific groups of PWE. Data handling was in accord with the Data Protection Act, 2018. Ethics approval was not required as all data was available to the public before initiation of the study.

## Results

There were a total of 2752 questions, all presented in the raw data format of natural language: The use of natural language resulted in duplication; therefore, an unsupervised topic modelling algorithm was utilised to process and categorise the data by topic into an aggregated question set. This acted as the first stage toward the supervised topic classification, with the additional benefit of providing a list of questions to be submitted for answering by healthcare professionals required for the Virtual Assistant.

The topic modelling reduced the questions to 163 topics. Questions were from PWE in the United States, UK, France, Germany and Australia. All data related to providers who operate only in the English language, impacting upon the countries reached and the topics of questions. This highlights the need for the Virtual Assistant to be able to process/translate to other languages to answer the concerns of all PWE worldwide, including those in remote areas.

The results of this exercise, both the analysis of the initial natural language questions and the topic modelled aggregate questions, provided insight not just into the questions PWE want answered but into the issues PWE are facing. As per Table 2, topics ranked highest focus upon aspects of daily life; the most asked questions being around the subject of keeping independence; both generally and in specific areas such as cooking/driving/exercise, and other social aspects of life with a focus on sexual relationships.

**Table 1.**
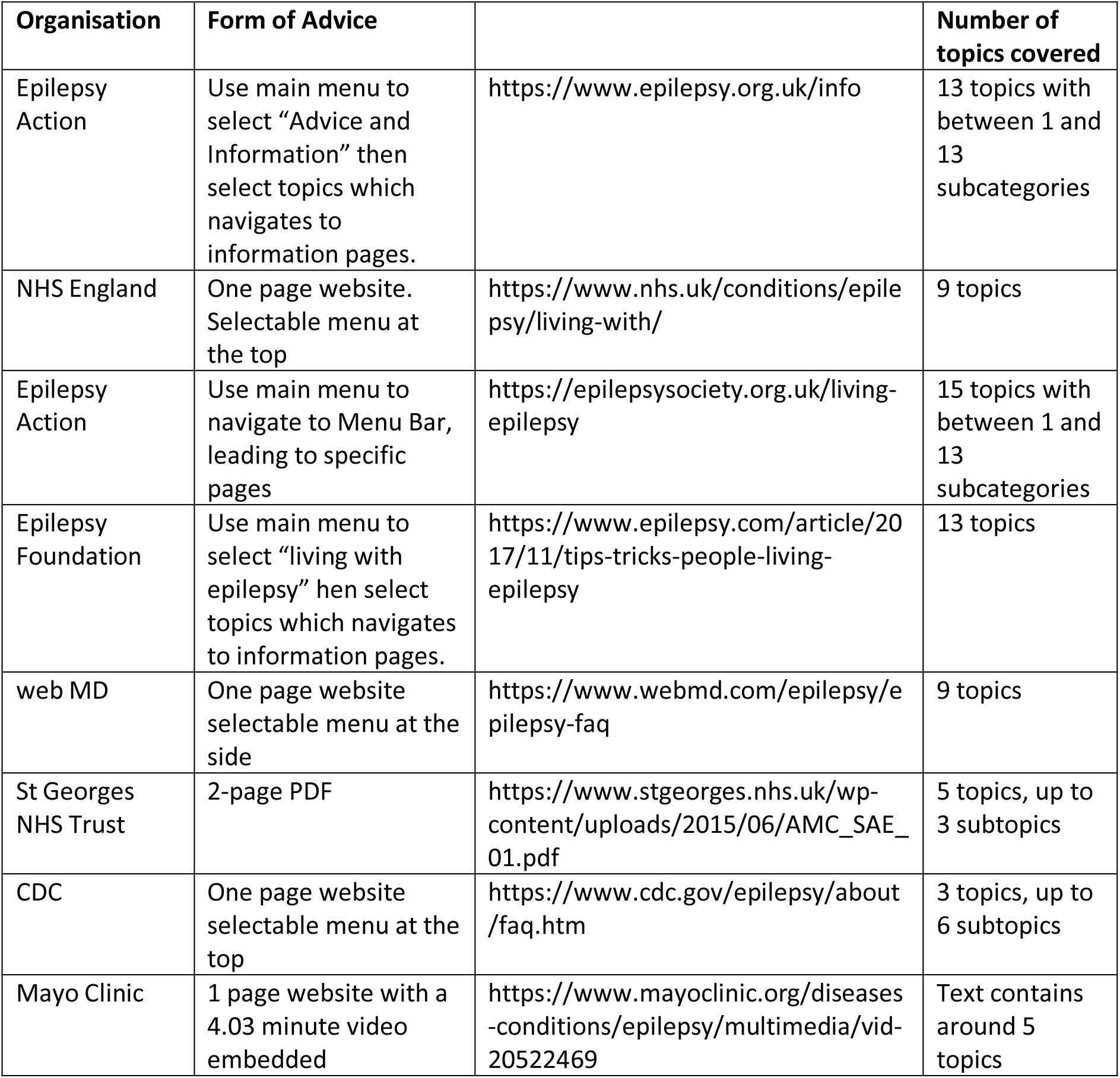
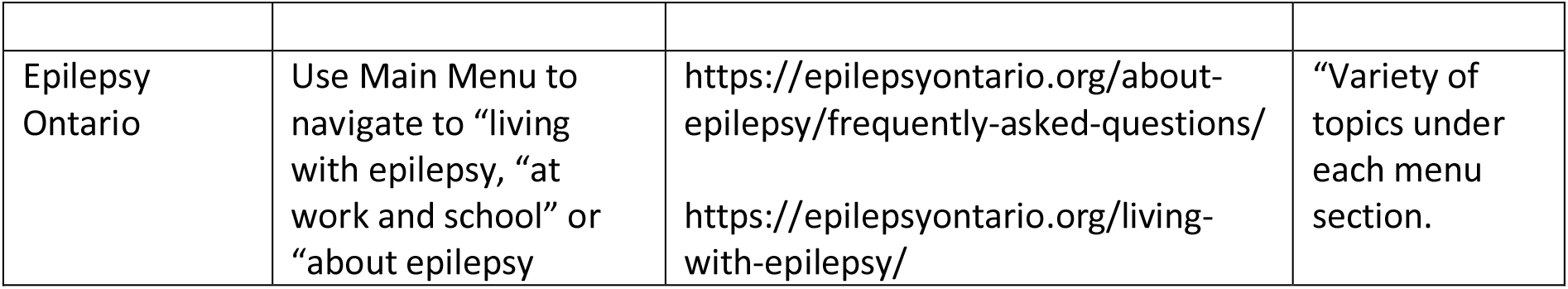
The table lists results from Google search

**Table 2.** Top 10 Aggregated Questions with Geographical Locations (for specific examples of questions see Supplementary data):

| Question (Aggregated) | Number of people<br>(% of total 2752) | UK | US<br>(% of Column 2) | CAN<br>(% of Column 2) | FRA<br>(% of Column 2) | GER<br>(% of Column 2) | AUS<br>(% of Column 2) |
| --- | --- | --- | --- | --- | --- | --- | --- |
| How do I keep my independence? | 84<br>(3%) | 61<br>(73%) | 19<br>(22.0%) |  |  |  | 4<br>(5%) |
| Is it normal to feel that epilepsy has ruined my life? | 77<br>(3%) | 52<br>(68%) | 23<br>(30.0%) | 1<br>(1.0%) |  |  | 1<br>(1%) |
| How do I know if I'm safe to exercise? | 62<br>(2%) | 52<br>(84%) | 9<br>(14%) |  |  |  | 1<br>(2%) |
| Do I have to take my medication? | 59<br>(2%) | 57<br>(96%) | 1<br>(2%) |  |  | 1<br>(2%) |  |
| How do I know what my seizure triggers are? | 57<br>(2%) | 49<br>(86%) | 8<br>(14%) |  |  |  |  |
| Will having sex cause seizures? | 55<br>(2%) | 48<br>(87%) | 7<br>(13%) |  |  |  |  |
| Can you have sex if you have epilepsy? | 54<br>(2%) | 52<br>(96%) | 2<br>(4%) |  |  |  |  |
| what is my risk of SUDEP? | 53<br>(2%) | 51<br>(96%) | 2<br>(4%) |  |  |  |  |
| What are the chances medication will control my seizures? | 51<br>(2%) | 38<br>(74%) | 1<br>(2%) | 5<br>(10%) |  |  | 7<br>(14%) |
| Can Covid trigger epilepsy or make it worse | 51<br>(2%) | 34<br>(67%) | 11<br>(21%) | 4<br>(8%) |  |  | 2<br>(4%) |
| Can I use knives/sharp or hot objects | 51<br>(2%) | 32<br>(63%) | 19<br>(37%) |  |  |  |  |

The overall data analysis suggests that the questions relating to general day to day living are asked by people from the UK (73.05%), whereas technical questions relating to different mitigations or treatments are asked by those from the US (80.85%). People in the UK (74.51%) asked more questions in the social media datasets reviewed (including US Platforms) than those in the US. It is unclear whether this is due to the difference in healthcare provision, or whether countries are better meeting patient needs? Questions relating to epilepsy and other conditions (for example menopause/ASD/stress) were equal in terms of the geographical location of origin.

The general tone of the language of questions was negative; focussing on what cannot be done, what is not permitted, the feeling that the PWE have lost control of their lives and concerns towards being perceived as “attention seeking”.

There was a high incidence of questions relating to when it was suitable to contact clinicians for advice – higher in the UK at 76%. There was obvious uncertainty as to what conversations with clinicians should concern. This raises the query as to whether basic questions are being answered by available literature and/or conversations with PWE/carers: are these questions expected by clinicians or does this suggest lack of understanding of the actual issues, where the focus less on the clinical aspects and more on the socio-economic? Conversely, does this evidence basic information being given by clinicians but not understood/retained by PWE at appointments?

Whilst medications and treatments are a consideration for PWE, questions focus on the side effects and need. Questions relating to specific treatment options (medication and alternatives) were more prevalent from the US than other countries (63%), indicating a possible difference between a National Health Service and private healthcare impacting upon the choice and cost benefit. The number of questions relating to the impact of COVID and vaccinations upon epilepsy indicate a lack of specific information for PWE.

Questions were asked by healthcare professionals (5%) demonstrating a lack of clarity amongst care services and a possible need for updated information for clinicians via such a virtual assistant – whether as a generic vehicle for PWE or a clinically specific app for clinicians to access correct, current advice. The dataset only utilised the status of “PWE, GP, Carer”; It would be interesting to investigate the breakdown of clinician and roles so that resources, services and advice can be tailored to provide support to these relevant groups, who may have very differing needs.

When read alongside the existing FAQs, the data indicates a mismatch between standard FAQ and patients concerns. Current resources do not provide enough information in terms of specificity and detail for PWE. PWE have concerns and questions which extend further than those that can be provided by a standard FAQ: more specific to the previous experiences/hopes and desires/acceptance of PWE than the standard FAQ can provide.

PWE expressed through their questions the desire for more opportunities to gather more detailed and clinical led information above and beyond the peer support model. This study indicates that technology can support this need. Technology and telehealth has the benefit (27) and drawback (28) for both staff and patients (29) of being anonymous, therefore there is value in further exploration into development of technological solutions combining peer support and clinical advice. There is a clear need for clinically correct answers and advice which is not possible due to time and financial constraints in the present clinician environment. A virtual assistant addresses this need; providing standardised information, in an understandable format, signposting to further detailed specific information.

## Discussion

This initial study indicates that further research, a deeper dive utilising innovative methods on an ongoing basis (e.g., social media, anonymised detailed feedback and analysis of the results of virtual assistants and other technology) would be useful to ensure services meet the wider, holistic need of PWE.

The next step is to develop a virtual assistant, initially in English, to act as a pilot. Assessment of a suitable platform adapted to the constantly evolving world of technology will be a starting point. With further extension this can become a place where PWE can access information, provide data for clinicians and researchers and assess the risks and impact of epilepsy upon their life at a given time: a “one stop shop” for clinicians, patients and carers. It is crucial that the platform used to build this enables not just future development but can analyse the trends and detail in real-time to ensure the service remains current and relevant. Such capability will increase the understanding of clinicians as to the impact of epilepsy upon the person behind the condition, the social rather than clinical model of care, increasing the accuracy with which resources and research can be targeted to meet needs.

Whilst the initial build will be in English, capabilities such as language translation will enable the service to be extended worldwide, to provide specific advice to areas where there is limited contact with specialists in epilepsy.

The initial assessment must consider the potential for extension for future developments (for example the ability to link with wearable technology). The system must not therefore be closed but can be developed to utilise the multitudinal, diverse and unforeseen ways that technology might expand (30): an expansion gathering momentum following the expedition in advancements to health technology as a response to the COVID crisis (31).

Evaluating the success of the pilot will be primarily one of data relating to usage. Detailed feedback will be required to ensure that exclusion is minimised: that the app does not become obsolete, is not easily accessible or attractive for use. Whilst there will always be an element of digital exclusion this must be minimised but not prevent the development of technology (32). Such feedback and research into patient interaction will inform as to the value, positives and negative of being to ask questions to an anonymous app.

## Data Availability

All data produced in the present study are available upon reasonable request to the authors.

https://www.myepilepsyteam.com/questions

https://brainablaze.com/comments/feed/

https://brainablaze.com/feed/

## Notes

### Competing Interest Statement

The authors have declared no competing interest.

### Funding Statement

This study did not receive any funding

### Author Declarations

No ethics approval was required because all of the data used was openly available to the public before the initiation of the study.

