## Supplementary Data Table 2 for "Mind the Gap! Developing Patient Responsive Information for Epilepsy"

| Question (Aggregated) | Number of people (% of total 2752) | UK | US (% of Column 2) | CAN (% of Column 2) | FRA (% of Column 2) | GER (% of Column 2) | AUS (% of Column 2) | Examples of individual questions |
|---|---|---|---|---|---|---|---|---|
| How do I keep my independence? | 84 (3%) | 61 (73%) | 19 (22.0%) | | | | 4 (5%) | Epilepsy seems to mean that people think they can constantly question me or run my life for me, is it the same for everyone? How much help do I really need? Do people have to go with me everywhere? What can I do not to rely on others for everything? I don't want someone watching while I have a bath, are there ways round this? How can I afford to still do what I want and not rely on everyone else for favours? I feel like I am being followed and watched with everything I do - am I paranoid? How do I tell people to let me do what I want without upsetting them? Why does everyone call me inspirational when I do anything, but also blame any illness/headache/time off work on the fact I am overdoing it and therefore shouldn't do anything? Do I have to be with my child all the time? What happens when my child grows and wants to go out - how do I keep them safe? Will my child need to live with me forever? If my child's epilepsy comes back, will they need to move back home? Can my child go to university and stay in halls? My mum has just been diagnosed with epilepsy; does she need to move in with me? I'm worried my dad will have a seizure and mum can't help him know they are older; how do I persuade them to move near us? |
| Is it normal to feel that epilepsy has ruined my life? | 77 (3%) | 52 (68%) | 23 (30.0%) | 1 (1.0%) | | | 1 (1%) | Everything has had to change - job, where I live, people are scared of seizures and me – can I do anything to change this? How do I get my life back? I feel as if epilepsy has ruined my life – is this normal? I just want my old life back, how do I do this? Can I ever get life the same as it was? Will life be changed forever? Am I being selfish for feeling as if I have lost everything? Everyone tells me that it could have been worse, but it doesn't feel that way - how do I tell them this? I keep being told I'm 'making too much of it' that it would be worse if I had cancer/dementia etc - but to me it doesn't feel like that, how do you get people to understand? Why does no one get that epilepsy is seizures but is also more than that? I keep being told I'm making too much of it, but at the same time friends have disappeared as they are scared, I might have a seizure - why is this? Will life ever be fun again? How do I balance having the life I want with seizures? Why are people scared to invite me out anymore - is there anything I can tell them, so I get a social life back? I've had to change job, where I live, lost my friends because of epilepsy - can I get any support? |
| How do I know if I'm safe to exercise? | 62 (2%) | 52 (84%) | 9 (14%) | | | | 1 (2%) | What exercise can I do? Are there limits as to how much? I only enjoy extreme sports and exercise - walking doesn't cut it – what can I do? How much is too much? If I exercise, will it cause a seizure? Is it usual to get seizures after exercise? Can I run alone? Can I swim alone? Can I do open water swimming? Can I surf? Can I rock climb? What do I tell people to do if I have a seizure while I'm exercising? Can I do cycle races? Can I race motorbikes because it's not on the road? Can I go on a bouncy castle? Can I trampoline? Can my child do gymnastics? What exercise can I advise patients to do? What are the limits to what my brother who has epilepsy can do? The meds make me feel so tired I can't be bothered and I'm getting fat, can I change meds so don't feel so lazy? Why does my doctor keep telling me to exercise? My son wants to play football, is this ok? Can rugby make epilepsy worse? |

| | | | | | | | | |
|---|---|---|---|---|---|---|---|---|
| Do I have to take my medication? | 59 (2%) | 57 (96%) | 1 (2%) | | | 1 (2%) | | I hate the meds - If the side effects are this bad why do I need to take them? I hate being ruled by meds - just pills and pills, can I get some I take once a day? If I get seizures anyway why bother taking the AEDs? If meds only reduce seizures not stop them is there a point to taking them? What do I do if I forget to take my meds? What do I do if I'm late taking my meds? Do I have to take my meds with water? How do I know if I'm on the best medication? How do doctors know what is the best medication? Are some medications safer than others? Do I get a choice of medications? Will medications damage my child's long-term prospects? |
| How do I know what my seizure triggers are? | 57 (2%) | 49 (86%) | 8 (14%) | | | | | Seizures happen - how do I know what is a trigger or not when it seems just life can trigger? How do I avoid all triggers when I can't do this and work? Is flashing lights always a trigger? Can sounds trigger seizures? Can music trigger seizures or is it the opposite? How do I find out my seizure triggers? Can certain types of food trigger seizures? What might trigger seizures for me? Can my GP help me work out what me triggers are? Does lack of sleep trigger seizures? Do I have to know what my triggers are - why does this help anyone? |
| Will having sex cause seizures? | 55 (2%) | 48 (87%) | 7 (13%) | | | | | Should I have sex? Will sex trigger seizures? What is the risk I will have a seizure during sex? Are certain sexual activities more likely to cause seizures than others? Are any sexual positions better than others to prevent seizures? Will having sex trigger a seizure? Are different sexual activities more likely to trigger seizures than others? Does the length of time we have sex for pose a risk to triggering seizures? How likely is it I will have a seizure during sex? |
| Can you have sex if you have epilepsy? | 54 (2%) | 52 (96%) | 2 (4%) | | | | | Will epilepsy frighten people from having sex with me? Can I still have sex? Does having epilepsy change the amount of sex I can have? Should I reduce how many times a week I have sex? My girlfriend has just been diagnosed with epilepsy; can she still have sex? If my wife has epilepsy, do I have to be gentler when having sex? Will it be safe to have oral sex in case my girlfriend has a seizure during? Will medications affect sex? Will my boyfriend still want to have sex with me? Is there a limit to the amount of sex we can have? We have different preferences for sex as enjoy BDSM, but my boyfriend has epilepsy - can we still do this or is it better to stop, I'm worried it might make his seizures worse? Is oral sex advisable (I have epilepsy, my boyfriend doesn't) or could it cause damage if I have a seizure? I'm gay - can we still have sex? What happens if I have a seizure during sex? Will my partner still want to have sex with me now - they are so worried? How do I talk to my partner about sex now I have epilepsy? Would psychosexual therapy be available? If I'm likely to have a seizure whilst having sex at which stage, is it likely to occur - arousal, during or orgasm? Can I have an orgasm without causing a seizure? Can you have multiple orgasms, or will this cause seizures? Can I masturbate? Can I use sex toys? |
| what is my risk of SUDEP? | 53 (2%) | 51 (96%) | 2 (4%) | | | | | How do I know if I am at risk of SUDEP? Will I be told what my SUDEP Risk is by my doctor? Do I need to worry about my SUDEP Risk? Should I know my SUDEP Risk? Apart from SUDEP does epilepsy shorten your life? |
| What are the chances medication will control my seizures? | 51 (2%) | 38 (74%) | 1 (2%) | 5 (10%) | | | 7 (14%) | I've tried several medications, and none are really stopping seizures. How much will medication control my seizures? Will medications stop my seizures? Will medications stop all seizures or just some? What does it mean if medications can't control my seizures? What do I do if medications don't control all my seizures? What type of seizures will medications stop? |
| Can Covid trigger epilepsy or make it worse | 51 (2%) | 34 (67%) | 11 (21%) | 4 (8%) | | | 2 (4%) | Will COVID make my seizures more frequent? Can COVID Cause epilepsy? Can COVID cause seizures? Does long COVID have an effect on seizure frequency? |

| | | | | | | | | |
|---|---|---|---|---|---|---|---|---|
| Can I use knives/sharp or hot objects | 51 (2%) | 32 (63%) | 19 (37%) | | | | | Am I allowed to use knives? Can I cook? Is there any way I can make cooking safer? Is there any better type of cooking device to use? Can I use a BBQ? Can I cook using gas? I enjoy cooking and don't want to have to give it up? Do I have to eat takeaways? If I can't cook and have to live on cold food or takeaways, how do I stop getting fat? Baking is my one love so what do I do if it means I have to stop? Can I use a knife when fishing or is this just cooking? I live on my own, so how do I cook? The charities say use an induction hob, but I can't afford one so how do I cook now? We have open fires, it's the only we can really afford to heat the house, what do I do as can't afford to stop using them? We go camping a lot (I'm a single mother) how do I cook, does this mean I can't go? |
